# Diagnostic accuracy of MRI in diagnosing Cardiac Sarcoidosis - A Meta-analysis

**DOI:** 10.1101/2023.04.29.23289313

**Authors:** Naqeeya Mustafa Sabuwala, Dwija Raval, Hetvi Parikh, Falak Saiyed, Dev Desai

## Abstract

**Introduction:** A multisystem inflammatory illness-sarcoidosis is characterized by organ failure, noncaseating granuloma development, and inflammation. The most frequently affected tissues are the lungs and pulmonary lymph nodes, although other tissues can also be affected including the heart. Cardiac Sarcoidosis can be lethal and result in conduction abnormalities, arrhythmias, and sudden cardiac death. As endocardial biopsy is invasive, Advanced imaging techniques, such as cardiac magnetic resonance (CMR) could be used to diagnose cardiac sarcoidosis and evaluate prognosis. Small and focal cardiac abnormalities have been successfully identified by (DE)-MRI. This comprehensive study and meta-analysis was carried out to aid in the CS diagnosis.

**Methodology:** Using a strategy based on the search terms-sarcoidosis and CMR/MRI separately, we searched-Pubmed, Google Scholar, Embose, Cochrane Library. Studies were disqualified if they lacked enough data to fill out a 2*2 contingency table. We strictly adhered to the Japanese criteria for diagnosing cases. Data from 12 studies with a total of 785 cases with TP of 212 was included.

**Results:** We found the cardiac MRI Sensitivity= 0.934 (95% CI = 0.904 to 0.964), Specificity = 0.875 (95% CI = 0.826 to 0.923), PPV= 0.752 (95% CI = 0.682 to 0.822), Younden index= 0.808 and AUC (Area under curve) for ROC plot= 0.904 with a Diagnostic accuracy of 0.892 for detecting sarcoidosis.

**Conclusion:** Cardiac MRI is a good and reliable screening and diagnostic tool that can be employed as a non-interventional method for diagnosis of Cardiac Sarcoidosis and future prognosis. Diagnostic accuracy of cardiac sarcoidosis using MRI, a meta-analysis.

## Introduction

Sarcoidosis is an inflammatory disease that affects many organs in the body, but mostly the lungs and lymph glands. In patients suffering from sarcoidosis, abnormal masses or nodules, inflamed tissues form in certain organs of the body. ^1^. Other organs are also involved, one of them being the heart. Cardiac involvement in patients with sarcoidosis is being recognized widely and is associated with poor prognosis.^2^ Initially uncommon, now Asians have recently been reported to have significant rates of cardiac involvement and unfavorable outcomes with 5-year mortality rates for cardiac sarcoidosis (CS) ranging from 25-67%.

The clinical symptoms of cardiac sarcoidosis (CS) include heart block, atrial arrhythmias, ventricular arrhythmias, and heart failure. The diagnosis of CS can be tough due to patchy infiltration of the myocardium. ^3^ Early options for treatment include corticosteroids or other immunosuppressive agents. Other treatments that can be used are placement of a pacemaker or implantable defibrillator to prevent sudden death.^4^

Cardiac involvement in sarcoidosis is difficult to diagnose clinically. A positive endomyocardial biopsy can help in the diagnosis of cardiac sarcoidosis but the sensitivity of biopsy is low, hence advanced imaging techniques, such as cardiac magnetic resonance (CMR) could be used to diagnose cardiac sarcoidosis and determine prognosis. Currently, CMR is recommended as the first-line imaging study for the diagnosis of CS, as it has a high negative predictive value. ^5^

The patient’s risk of getting exposed to ionizing radiation is very low, and myocardial MRI has a good spatial resolution. Small and focal cardiac abnormalities have been successfully identified by (DE)-MRI.

This comprehensive study and meta-analysis was carried out to aid in the CS diagnosis.

## Methodology

### Data collection

Using a strategy based on the search terms-sarcoidosis and CMR/MRI separately we searched -Pubmed, GoogleScholar, Embose, Cochrane Library. This systematic review was conducted following the Preferred Reporting Items for Systematic Reviews and Meta-Analyses (PRISMA) statement.

### Inclusion and Exclusion Criteria

Studies were considered if they (1) evaluated the CMR’s diagnostic efficacy for sarcoidosis, and (2) used Japanese diagnostic criteria. Studies were disregarded if they did not assess cardiac sarcoidosis using CMR or if there was inadequate information in them.

We preferred studies that had biopsy-proven diagnoses as a comparison over others.

### Data Extraction and Quality Assessment

Two reviewers independently retrieved and carefully screened the abstracts of articles to identify relevant articles. The same two reviewers independently collected data from the primary studies including publication details (author name and year of publication), number of patients, and then the Sensitivity, Specificity, Positive Predictive Value (PPV), Area Under the Curve of Receiver Operating Characteristic Curve (AUC of ROC) and Youden’s Index (YI) of DECT as diagnostic tools were calculated.

We didn’t limit the searches to specific study methodologies or publication dates. We used a technique based on the search terms (sarcoidosis and magnetic resonance imaging) separately to conduct a search from January 1, 1980, to January 18, 2023, and we included all research on the reliability of diagnostic tests. Studies that lacked the data necessary to fill out a 2*2 table were disregarded. We incorporated the most recent findings from studies conducted on the same population.

### Statistical analysis

Analysis of data was performed using Excel, RevMan (Review Manager, version 5.33), SPSS (Statistical Package for Social Sciences), and Stata 14.0. The presence of bias in the included studies was evaluated by Quadas-2. Individual study sensitivity and specificity were plotted on forest plots and in the receiver operating characteristic (ROC) curve. ^6^We calculated the summary positive and negative likelihood ratios using the Fagan plot analysis command strata 14.0.

## RESULTS

PubMed database, Google Scholar search, Embose, and the Cochrane Library were used for the collection of 966 articles. After reviewing the titles, abstracts, study types, and full texts and excluding duplicate and unrelated articles, twelve studies were included in this meta-analysis. A total of 785 patients with CS were available for the meta-analysis. The main information and characteristics of the included studies are summarised in Table 1. Cardiac MRI has an overall sensitivity of 0.934 (95% CI = 0.904 to 0.964) and specificity of 0.875 (95% CI = 0.826 to 0.923) in the diagnosis of CS on forest plot (Figure 1). The summary of the ROC curve is given in Figure 2. The area under the curve was 0.904 with a Diagnostic accuracy of 0.892 for detecting sarcoidosis. The overall diagnostic odds ratio was 98.530. The Fagan plot analysis is shown in Figure 3. It showed a pretest probability of .29; a positive likelihood ratio of 7.44; A positive posterior probability of .75; a negative likelihood ratio, of 0.008; and the negative posterior probability of .03. The Youden index is 0.808. The diagnostic accuracy is 0.892. Figure 4 and Table 2 is bias study of the meta-analysis. The tool used for this is QUADAS-2 analysis.

**FIGURE 1:**
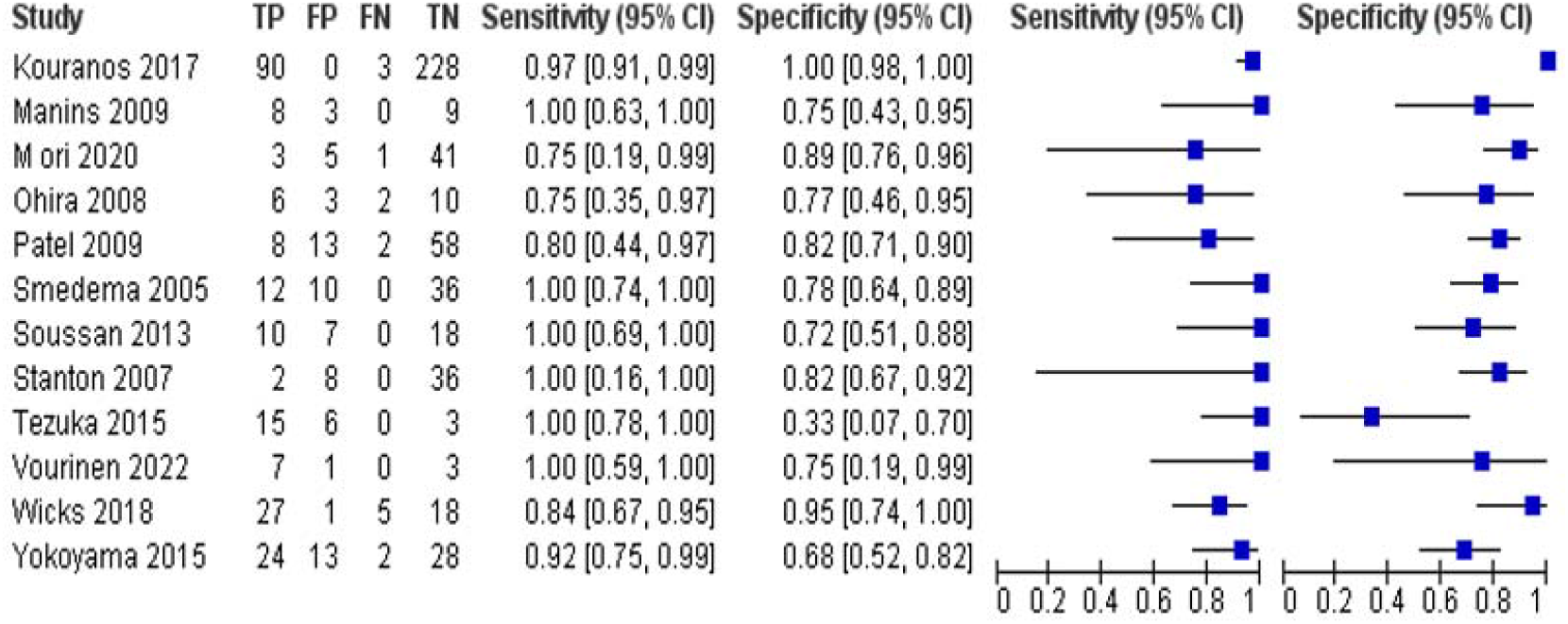
Forest plots of sensitivity and specificity Cardiac MRI has an overall sensitivity of 0.934 (95% CI = 0.904 to 0.964) and specificity of 0.875 (95% CI = 0.826 to 0.923) in the diagnosis of CS.

**TABLE 1:** Characteristics of the study

| STUDY | YEAR | NO.OF PATIENTS | SENSITIVITY | SPECIFICITY | TP | FP | FN | TN |
| --- | --- | --- | --- | --- | --- | --- | --- | --- |
| **Kouranos**<sup>7</sup> | 2017 | 321 | 0.97 | 1 | 90 | 0 | 3 | 228 |
| **Manins**<sup>8</sup> | 2009 | 20 | 1.00 | 0.75 | 8 | 3 | 0 | 9 |
| **Mori**<sup>9</sup> | 2020 | 50 | 0.75 | 0.89 | 3 | 5 | 1 | 41 |
| **Ohira**<sup>10</sup> | 2008 | 21 | 0.75 | 0.77 | 6 | 3 | 2 | 10 |
| **Patel**<sup>11</sup> | 2009 | 81 | 0.80 | 0.82 | 8 | 13 | 2 | 58 |
| **Smedema**<sup>12</sup> | 2005 | 58 | 1.00 | 0.78 | 12 | 10 | 0 | 36 |
| **Soussan**<sup>13</sup> | 2013 | 35 | 1.00 | 0.72 | 10 | 7 | 0 | 18 |
| **Stanton**<sup>14</sup> | 2007 | 46 | 1.00 | 0.82 | 2 | 8 | 0 | 36 |
| **Tezuka**<sup>15</sup> | 2015 | 24 | 1.00 | 0.33 | 15 | 6 | 0 | 3 |
| **Vourinen**<sup>16</sup> | 2022 | 11 | 1.00 | 0.75 | 7 | 1 | 0 | 3 |
| **Wicks**<sup>17</sup> | 2018 | 51 | 0.84 | 0.95 | 27 | 1 | 5 | 18 |
| **Yokoyama**<br>18 | 2015 | 67 | 0.92 | 0.68 | 24 | 13 | 2 | 18 |

**Table 2:**
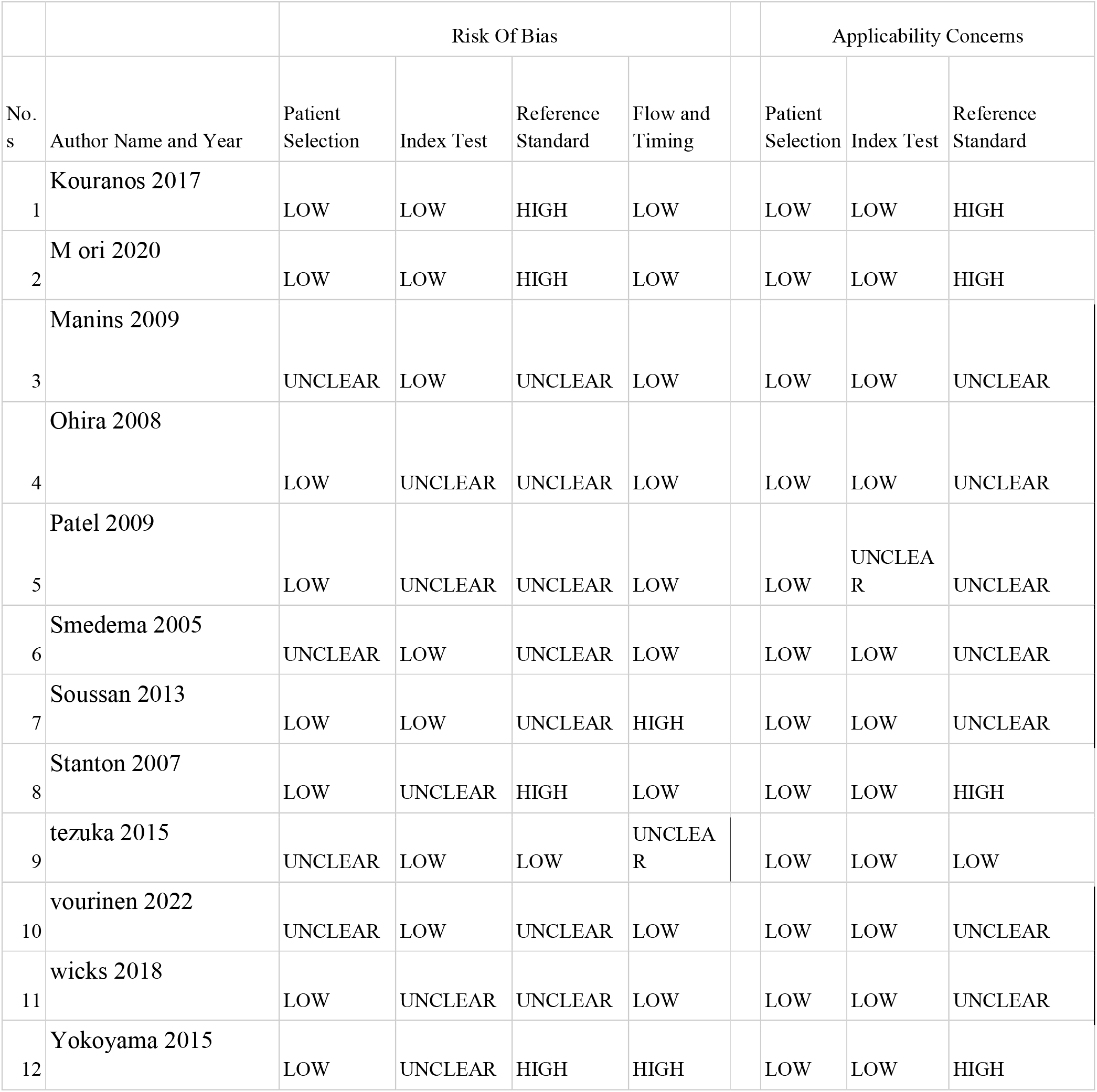
Bias Study by QUADAS-2 Analysis

**FIGURE 2:**
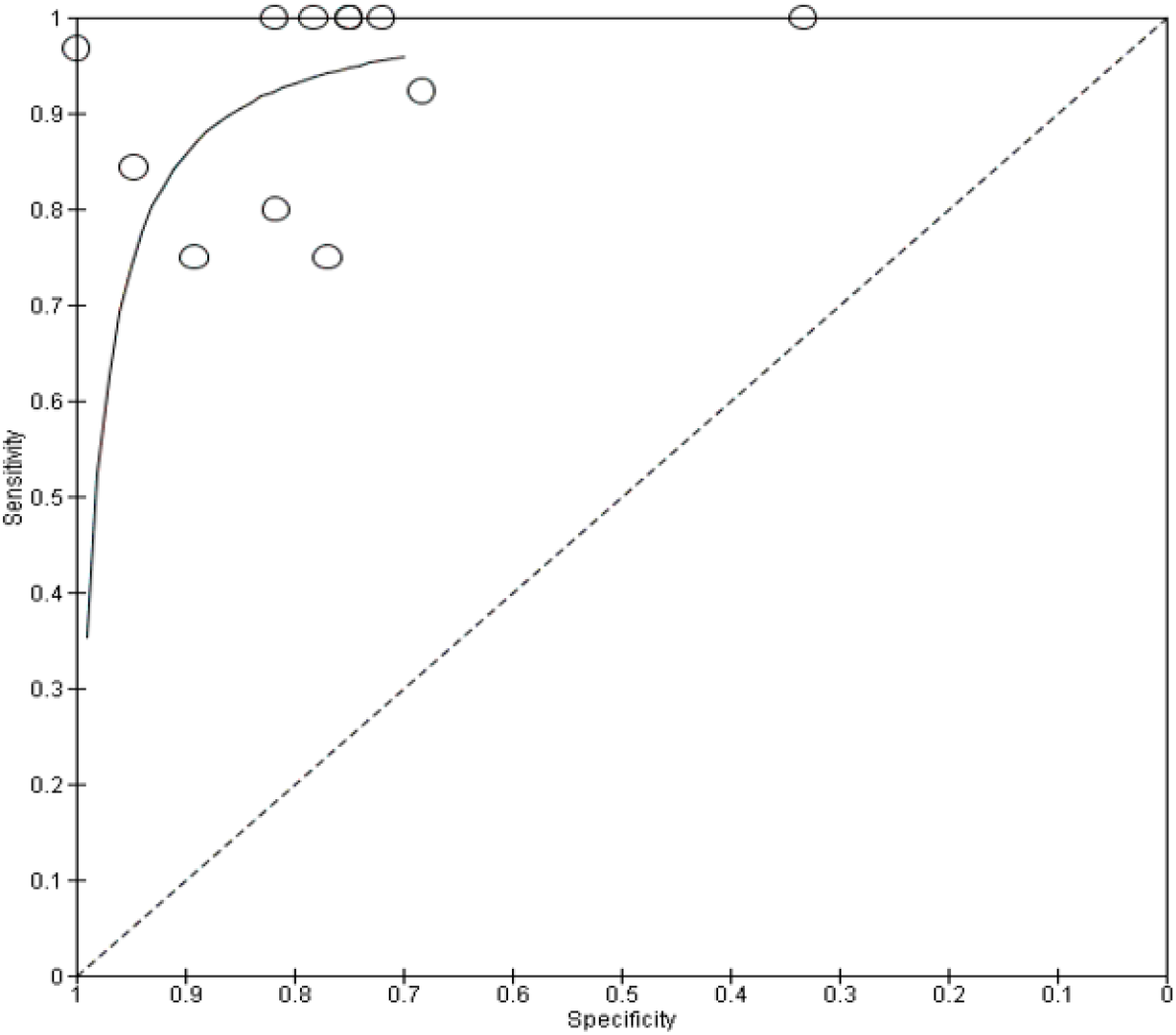
The summary of the receiver operating characteristic (ROC) curve. The area under the curve was 0.904. The overall diagnostic odds ratio was 98.530.

**FIGURE 3:**
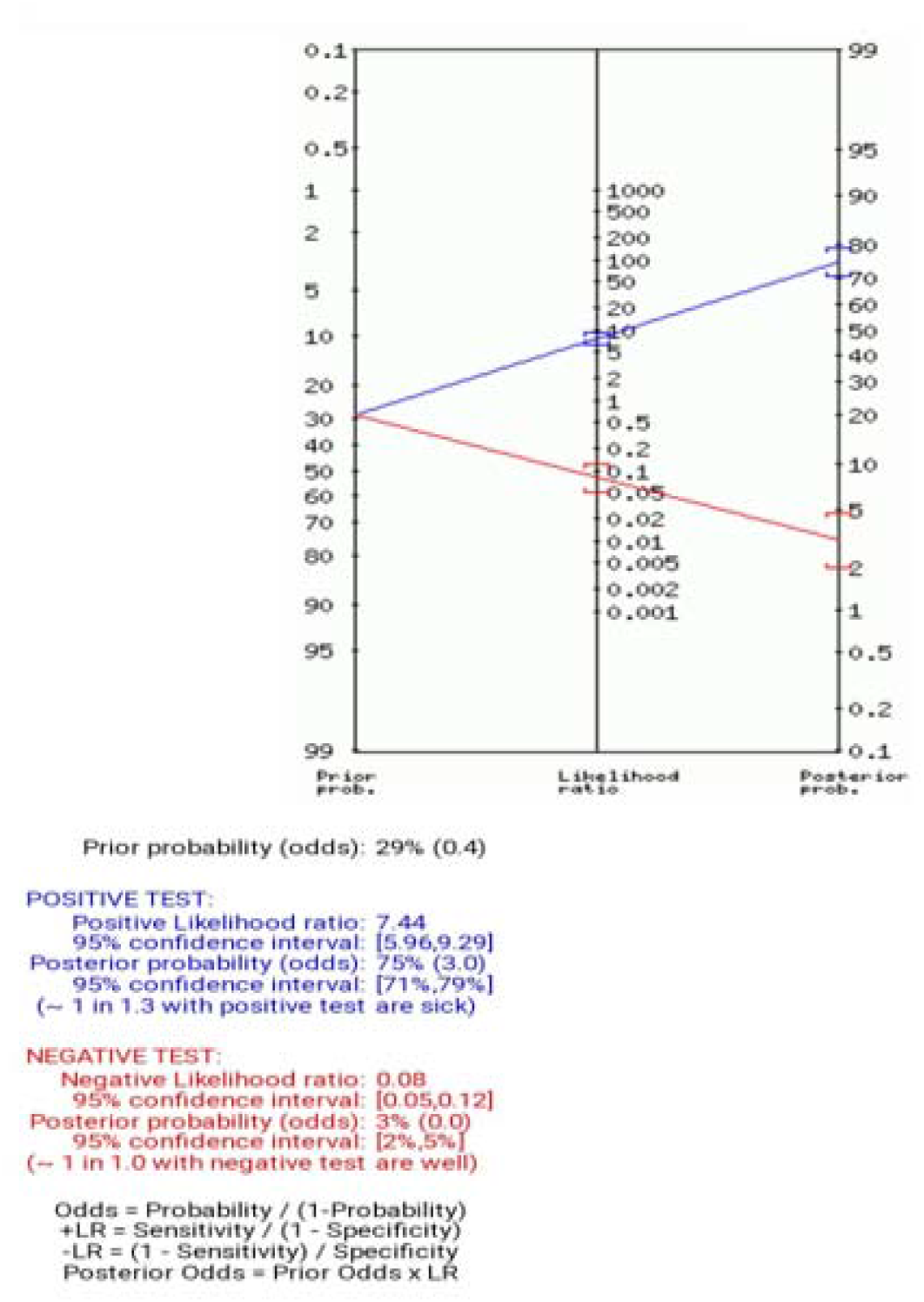
Fagan Plot analysis. pretest probability .29; positive likelihood ratio 7.44; A positive posterior probability of .75; negative likelihood ratio, 0.008; and the negative posterior probability .of .03

**Figure 4:**
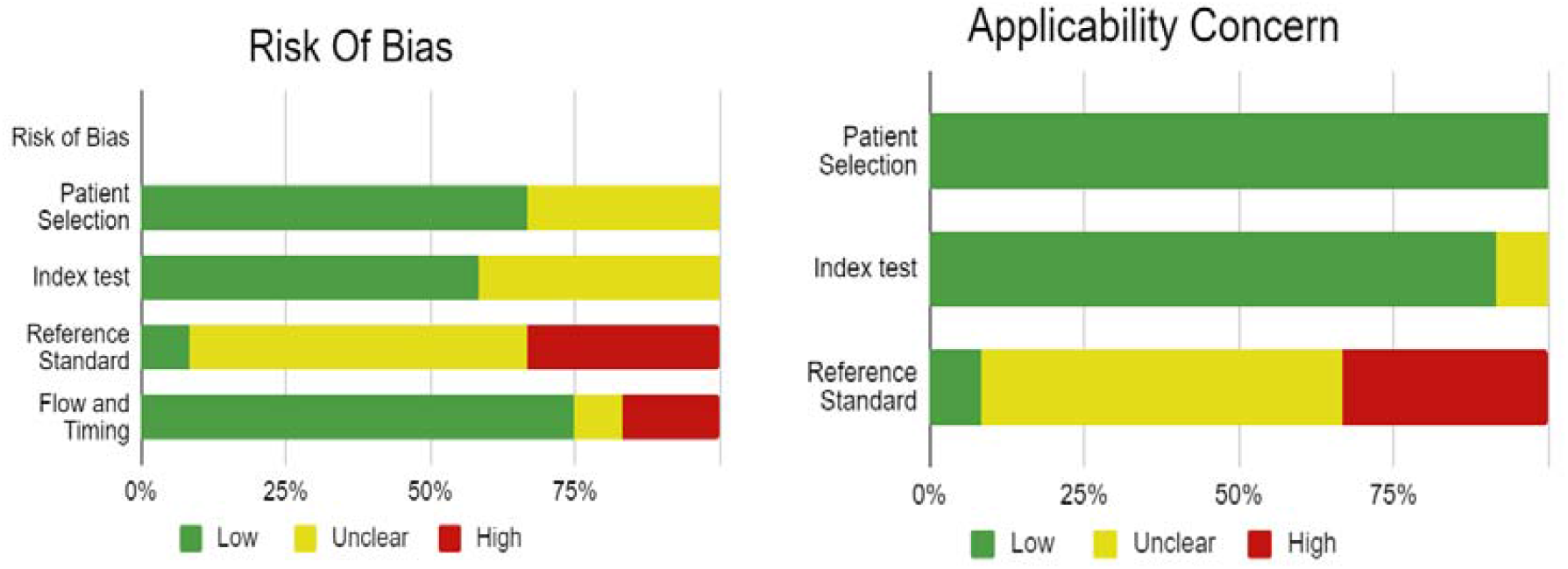
Bias study by QUADAS-2 analysis

**Fig. 5:**
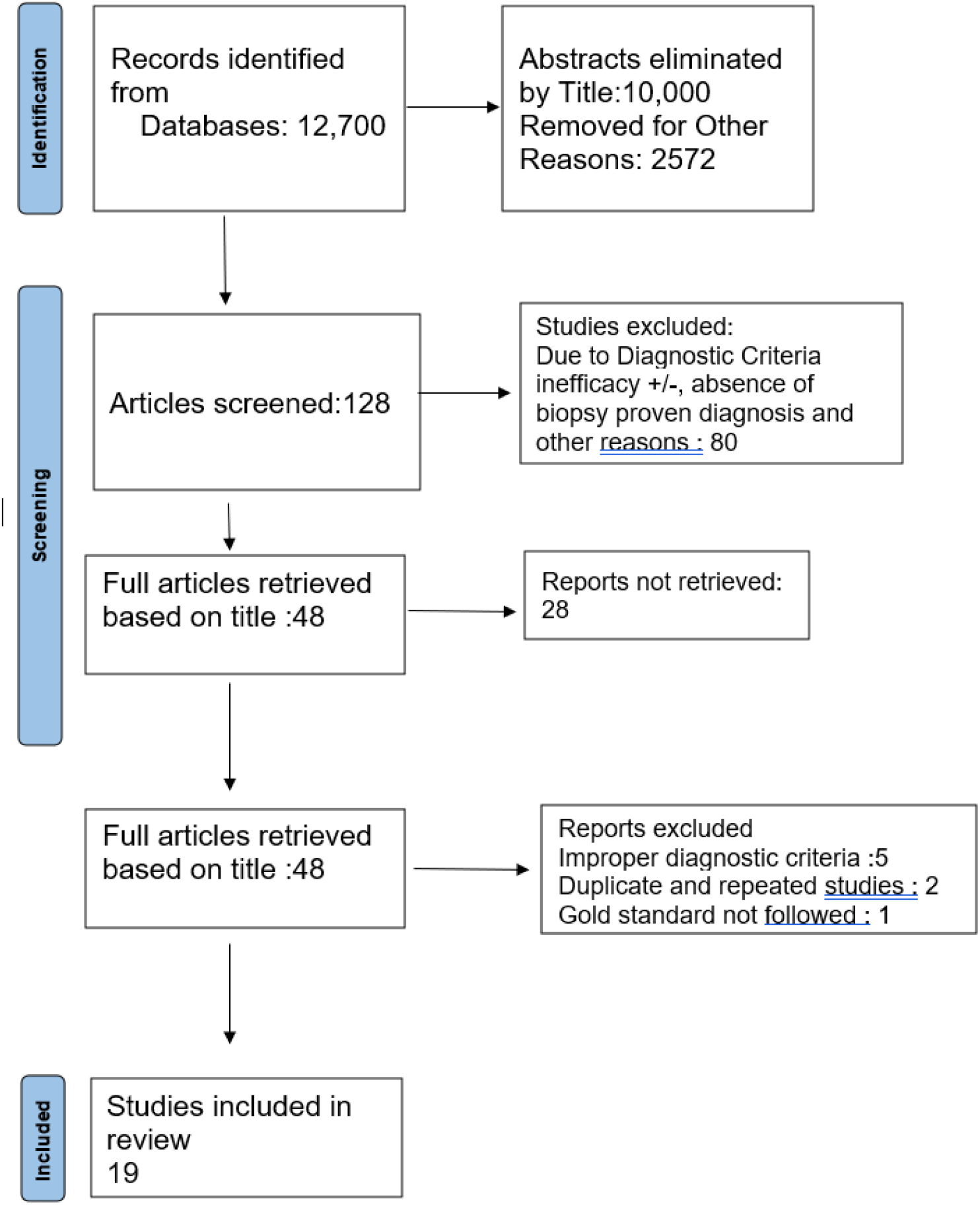
PRISMA CHART.

## Discussion

Sarcoidosis is an inflammatory disorder that affects multiple organs. In cardiac sarcoidosis, collections of immune cells form granulomas in the heart. This can result in arrhythmias, such as ventricular tachycardia or heart block. In this study, cardiac magnetic resonance (CMR) is used to diagnose cardiac sarcoidosis and evaluate prognosis. Fourteen studies were included in this study. These studies have different values of sensitivity and specificity. According to Kim ^11^, in consecutive patients with biopsy-proven extracardiac sarcoidosis who were prospectively screened for cardiac involvement, the principal finding was that DE-CMR identified myocardial abnormalities in significantly more patients than a standard clinical evaluation. They found that DE-CMR is more than twice as sensitive for cardiac involvement as current consensus criteria in patients with sarcoidosis. Smedema ^12^. evaluated the diagnostic role of cardiac MRI in 58 patients with suspected CS and reported that DE showed favorable sensitivity and specificity of 100% and 78%, respectively.

According to a study done by Matthew ^19^, cardiac MRI and FDG PET had sensitivities of 92% and 81% and specificities of 72% and 82%, respectively, implicating that Cardiac MRI has a higher sensitivity but similar specificity than fluorodeoxyglucose PET. According to Jianxiong ^20^, CMR had an overall sensitivity of 0.93 (95% CI, 0.87–0.97) and specificity of 0.85 (95% CI, 0.68– 0.94) in the diagnosis of cardiac sarcoidosis. They also stated that CMR holds promise for the detection of regional interstitial edema and scarring consistent with CS.

Wicks ^17^ suggested that the presence of LGE and FDG uptake on hybrid cardiac PET/MR identifies patients at higher risk of death, arrhythmia, and decompensated heart failure and should be considered in the assessment of all patients presenting with suspected CS to determine disease activity and prognosis. According to Shimada ^21^, images of the heart obtained by Gd-MRI may reflect active inflammation with interstitial edema in patients with sarcoidosis. Gd-MRI may be a useful noninvasive method for the early detection of cardiac sarcoidosis and for evaluating the effects of steroid therapy.

However, as per Sharma ^22^, CMR has currently one limitation: it cannot be performed in patients with CS who carry cardiac devices, such as cardiac defibrillators (AICD) or pacemakers, which are contraindications to CMR. Another limitation as stated by Isobe ^23^, is the inability to use gadolinium to image patients with renal impairment.

The clinical presentation of Cardiac sarcoidosis may range from an accidental discovery to Heart failure, various arrhythmias, and sudden death. Hence, in addition to Cardiac MRI, various other methods like electrocardiography, 24-h Holter monitoring, echocardiography, FDG-PET, etc are used to identify this condition as early as possible. Tomas Vita ^24^ concluded that among patients with suspected CS, combining CMR and PET provides complementary value for estimating the likelihood of CS and guiding patient management. Furthermore, many studies are needed to determine the diagnostic accuracy of Cardiac MRI in patients with cardiac sarcoidosis.

## Conclusion

As a result of this meta-analysis, it is possible to diagnose cardiac sarcoidosis and screen people who may have the disease. The diagnostic efficacy of MRI increased along with the technique’s advancement. It is essential to thoroughly test the patients when cardiac involvement is suspected. Corticosteroid medication should be started as soon as possible to reduce negative effects.

We suggest, based on these findings, that non-invasive imaging modalities be used to diagnose iCS without histologic evidence in order to prevent the underdiagnosis of cases that may be treated. The clinical definition of iCS requires additional assessment, which is required.

## Data Availability

All data produced in the present work are contained in the manuscript

